# Prevalence and duration of detectable SARS-CoV-2 nucleocapsid antibody in staff and residents of long-term care facilities over the first year of the pandemic (VIVALDI study): prospective cohort study

**DOI:** 10.1101/2021.09.27.21264166

**Authors:** Maria Krutikov, Tom Palmer, Gokhan Tut, Christopher Fuller, Borscha Azmi, Rebecca Giddings, Madhumita Shrotri, Nayandeep Kaur, Panagiota Sylla, Tara Lancaster, Aidan Irwin-Singer, Andrew Hayward, Paul Moss, Andrew Copas, Laura Shallcross

## Abstract

**Background:** Long Term Care Facilities (LTCF) have reported high SARS-CoV-2 infection rates and related mortality, but the proportion infected amongst survivors and duration of the antibody response to natural infection is unknown. We determined the prevalence and stability of nucleocapsid antibodies – the standard assay for detection of prior infection - in staff and residents from 201 LTCFs.

**Methods:** Prospective cohort study of residents aged >65 years and staff of LTCFs in England (11 June 2020-7 May 2021). Serial blood samples were tested for IgG antibodies against SARS-CoV-2 nucleocapsid protein. Prevalence and cumulative incidence of antibody-positivity were weighted to the LTCF population. Cumulative incidence of sero-reversion was estimated from Kaplan-Meier curves.

**Results:** 9488 samples were included, 8636 (91%) of which could be individually-linked to 1434 residents or 3288 staff members. The cumulative incidence of nucleocapsid seropositivity was 35% (95% CI: 30-40%) in residents and 26% (95% CI: 23-30%) in staff over 11 months. The incidence rate of loss of antibodies (sero-reversion) was 2·1 per 1000 person-days at risk, and median time to reversion was around 8 months.

**Interpretation:** At least one-quarter of staff and one-third of surviving residents were infected during the first two pandemic waves. Nucleocapsid-specific antibodies often become undetectable within the first year following infection which is likely to lead to marked underestimation of the true proportion of those with prior infection. Since natural infection may act to boost vaccine responses, better assays to identify natural infection should be developed.

**Funding:** UK Government Department of Health and Social Care.

**Research in context:** *Evidence before this study:* A search was conducted of Ovid MEDLINE and MedRxiv on 21 July 2021 to identify studies conducted in long term care facilities (LTCF) that described seroprevalence using the terms “COVID-19” or “SARS-CoV-2” and “nursing home” or “care home” or “residential” or “long term care facility” and “antibody” or “serology” without date or language restrictions. One meta-analysis was identified, published before the introduction of vaccination, that included 2 studies with a sample size of 291 which estimated seroprevalence as 59% in LTCF residents. There were 28 seroprevalence surveys of naturally-acquired SARS-CoV-2 antibodies in LTCFs; 16 were conducted in response to outbreaks and 12 conducted in care homes without known outbreaks. 16 studies included more than 1 LTCF and all were conducted in Autumn 2020 after the first wave of infection but prior to subsequent peaks. Seroprevalence studies conducted following a LTCF outbreak were biased towards positivity as the included population was known to have been previously infected. In the 12 studies that were conducted outside of known outbreaks, seroprevalence varied significantly according to local prevalence of infection. The largest of these was a cross-sectional study conducted in 9,000 residents and 10,000 staff from 362 LTCFs in Madrid, which estimated seroprevalence in staff as 31·5% and 55·4% in residents. However, as this study was performed in one city, it may not be generalisable to the whole of Spain and sequential sampling was not performed. Of the 28 studies, 9 undertook longitudinal sampling for a maximum of four months although three of these reported from the same cohort of LTCFs in London. None of the studies reported on antibody waning amongst the whole resident population.

*Added value of this study:* We estimated the proportion of care home staff and residents with evidence of SARS-CoV-2 natural infection using data from over 3,000 staff and 1,500 residents in 201 geographically dispersed LTCFs in England. Population selection was independent of outbreak history and the sample is therefore more reflective of the population who reside and work in LTCFs. Our estimates of the proportion of residents with prior natural infection are substantially higher than estimates based on population-wide PCR testing, due to limited testing coverage at the start of the pandemic. 1361 individuals had at least one positive antibody test and participants were followed for up to 11 months, which allowed modelling of the time to loss of antibody in over 600 individuals in whom the date of primary infection could be reliably estimated. This is the longest reported serological follow up in a population of LTCF residents, a group who are known to be most at risk of severe outcomes following infection with SARS-CoV-2 and provides important evidence on the duration that nucleocapsid antibodies remained detectable over the first and second waves of the pandemic.

*Implications of all available research:* A substantial proportion of the LTCF population will have some level of natural immunity to infection as a result of past infection. Immunological studies have highlighted greater antibody responses to vaccination in seropositive individuals, so vaccine efficacy in this population may be affected by this large pool of individuals who have survived past infection. In addition, although the presence of nucleocapsid-specific antibodies is generally considered as the standard marker for prior infection, we find that antibody waning is such that up to 50% of people will lose detectable antibody responses within eight months. Individual prior natural infection history is critical to assess the impact of factors such as vaccine response or protection against re-infection. These findings may have implications for duration of immunity following natural infection and indicate that alternative assays for prior infection should be developed.

## Introduction

Older people in Long Term Care Facilities (LTCFs) have experienced the greatest burden of deaths from severe acute respiratory syndrome coronavirus-2 (SARS-CoV-2) infection. Contributing factors include immune-senescence and age-mediated inflammatory immune responses which increase susceptibility to severe infection together with close proximity to infected individuals within the enclosed facility. ^1^ Approximately 410,000 older people live in 11,000 LTCFs in England ^2^ but, partly due to limited PCR testing at the start of the pandemic, the true burden of SARS-CoV-2 infection in this population is unknown. Antibodies against viral proteins, including the nucleocapsid and the spike proteins, are produced in response to SARS-CoV-2 infection ^3^ and are likely to correlate with immunity against reinfection. ^4^ There is also growing evidence that previously infected individuals develop greater antibody responses to SARS-CoV-2 vaccination. ^5–7^ Robust antibody production has been demonstrated in older LTCF residents who have survived SARS-CoV-2 infection, ^8–10^ however large-scale sero-surveys to describe the extent of past exposure and immunity in this population are lacking.

There have been three waves of infection since the start of the COVID-19 pandemic in the UK (wave 1: March-May 2020; wave 2: September 2020-April 2021, wave 3: May 2021-curent) with most deaths in LTCF residents occurring in the first wave. ^11,12^ To curb the impact of the pandemic in vulnerable populations, widespread vaccination was rolled out in LTCFs from 8 December 2020 resulting in a decline in infections and deaths. ^12,13^ As the spike protein is the only immunogen present within most current vaccines, detection of antibodies against viral nucleocapsid can be used as an indicator of prior natural infection.^14,15^

Waning antibody titres in the months following natural infection have been observed across different populations. ^16,17^ Nucleocapsid-specific antibodies decline more rapidly than spike antibodies although the clinical significance of this is still unknown. ^18,19^ This is a potential concern as many research and surveillance studies use antibodies to nucleocapsid to differentiate patients with and without prior natural infection in order to estimate vaccine effectiveness. As such, definitive information on the prevalence and stability of nucleocapsid-specific antibodies within specific populations is required. We conducted a prospective seroprevalence survey in 201 LTCFs in England in order to estimate the proportion of surviving staff and residents in LTCFs with antibodies to nucleocapsid protein and the duration of antibody-positivity in this group.

## Methods

### Study design and participants

From June 2020, the VIVALDI study has been prospectively recruiting staff and residents from 201 geographically dispersed LTCFs in England which provide care to adults aged over 65 years.^20^ LTCFs vary in size and include a mix of “For Profit” and “Not for Profit” Providers.

In brief, eligible LTCFs were identified by the Provider or the National Institute for Health (NIHR) Clinical Research Network (CRN). Written informed consent was obtained from individuals in these LTCFs and where individuals were assessed as lacking capacity to consent, a nominated or personal consultee acted on their behalf. Sequential rounds of blood sampling were undertaken at eight-week intervals for the first two to three rounds of sampling, while subsequent rounds were set to follow first and second dose vaccine administration. Individuals donated a maximum of four samples between 11 June 2020 and 7 May 2021 and were able to join the study at any round.

LTCF staff and residents underwent regular screening (weekly in staff, monthly in residents) to detect SARS-CoV-2 viral RNA using polymerase chain reaction (PCR) testing of nasopharyngeal swabs, and additional testing as part of outbreak investigations.^21^ We excluded staff aged >65 years and residents <65 years. Where role was unknown, an age threshold of 65 was used to differentiate between staff and residents.

#### Data linkage

Participants were linked to a unique pseudo-identifier based on their National Health Service (NHS) identification number. Antibody results were submitted to NHS England (NHSE) and linked to PCR test results using the unique pseudo-identifier in the NHS COVID-19 Datastore, a secure online data repository established in response to the pandemic (https://data.england.nhs.uk/covid-19/). Every care facility in the UK is allocated a unique identifier by the Care Quality Commission (CQC), the national social care regulatory body. Individual-level records on diagnostic ICD-10 admission codes and dates of hospitalisation were retrieved from the Hospital Episode Statistics (HES) database which is maintained by NHSE and linked within the COVID-19 datastore by pseudo-identifier. Maximum monthly bed occupancy and staffing numbers were retrieved from the Capacity Tracker dataset (https://www.necsu.nhs.uk/capacity-tracker), a tool completed by LTCFs that collects operational data.

#### Study procedures

All serum samples were tested for IgG to nucleocapsid protein using the semi-quantitative Abbott ARCHITECT i-system (Abbott, Maidenhead) immunoassay. An index value of 0.8 was defined as positive (>=0.8) to maximise test sensitivity whilst maintaining specificity. ^18,19^ Quantitative IgG titres were obtained using the V-PLEX COVID-19 respiratory panel 2 kit (96-well, 10 Spot Plate coated with SARS CoV-2 antigens [spike protein, spike receptor-binding domain]; K15372U; Meso Scale Diagnostics, Rockville, MD, USA). The positivity cut-offs were determined by the testing laboratory. ^24^

RT-PCR testing of nasopharyngeal swabs was conducted in a national network of laboratories, established as part of the pandemic response.

##### Weighted Seroprevalence

All samples from eligible LTCFs were included in the seroprevalence analysis.

##### Time to antibody-loss

Time to antibody-loss was estimated in individuals that who could be linked to a pseudo-identifier and had at least two eligible samples separated by more than four weeks. It was not possible to ascertain the precise date of infection for most participants due to limited PCR testing. As such, individuals were included in the analysis if the date of infection and therefore seroconversion could be inferred through:

a. PCR-positive test prior to first positive antibody test: seroconversion was assumed to occur 14 days following the PCR date. (n=108)
b. Hospitalisation with confirmed or suspected COVID-19 prior to first positive antibody test: seroconversion was the admission date. (n=5)
c. A negative antibody test that preceded a positive antibody test: seroconversion date was taken to be the mid-point between tests. (n=3)
d. First positive antibody test was before 1 August 2020 therefore primary infection occurred during the first pandemic wave: seroconversion was taken as 5 May 2020 (14 days after the peak of cases nationally in wave 1). ^12^ (n=403)

Where it was possible to estimate seroconversion dates based on more than one criterion, these were prioritised in the following order; PCR date (a), hospitalisation (b) and seroconversion (c;d).

Individuals in whom the estimated date of seroconversion was > 120 days before the first antibody test were excluded to improve the representativeness of our sample. Those with an interval >120 days are likely unrepresentative of all individuals that initially developed antibodies in our cohort as there will be additional individuals who sero-reverted prior to their first antibody test.

##### Sero-reverted cohort

Samples from individuals who sero-reverted based on the Abbott assay result underwent additional testing to detect antibodies against spike and RBD.

Ethical approval for this study was obtained from the South Central—Hampshire B Research Ethics Committee (20/SC/0238).

### Statistical analysis

Unweighted estimates of seropositivity were calculated for all samples according to region, type of care provider, LTCF size, and testing interval. We also estimated the proportion of individuals with at least one positive sample over the sampling period. LTCF level estimates of seroprevalence were calculated by testing round.

#### Weighted seroprevalence and ‘cumulative incidence’

Weighted seroprevalence estimates were calculated separately for staff and residents and stratified by testing interval (1: June-July 2020; 2: August-September 2020; 3: October-November 2020; 4: December 2020-January 2021; 5: February 2021; 6: March-April 2021). These testing intervals were selected to be more representative of the study population, as they reflected the period of time that it took for most of the LTCFs in the study to undergo one round of testing. To account for LTCF-level testing rate, weights were calculated separately for staff and residents per sampling round using N/n, where N is the maximum bed occupancy or maximum total staffing for that month and n is the number of residents or staff tested. A ‘cumulative incidence’ that was weighted using N/n over the complete testing period in each LTCF was calculated overall and separately for staff and residents, defined as the proportion of individuals tested in the entire study period who tested positive at least once.

#### Time to antibody loss

We estimated the cumulative incidence of nucleocapsid-antibody waning in staff and residents separately by fitting Kaplan-Meier curves and compared using log-rank test. The event of interest was a negative antibody test (sero-reversion) following a positive antibody test. Time at risk was taken in days from the estimated date of sero-conversion, defined above. The date of sero-reversion was the mid-point between last antibody-positive and first antibody-negative test. Individuals who did not sero-revert (fail) were censored at the date of their last antibody test.

A sub-group analysis estimated cumulative incidence of sero-reversion by severity of primary infection, defined according to history of COVID-19 hospitalisation (confirmed or suspected).

#### Sero-reverted cohort

In the sero-reverted cohort, quantitative antibody titres were compared between staff and residents at 3 points; first positive antibody test, 30 days, and 60-90 days following estimated sero-reversion. Date of sero-reversion followed the definitions outlined above according to the Abbott assay.

Sample size for the VIVALDI study was originally based on the precision of estimates for antibody prevalence. However, the time horizon and number of included LTCFs have since expanded to address rapidly changing policy priorities. Therefore, no sample size calculations were done for this article. ^20^

A p-value <0·05 was considered to indicate statistical significance.

All statistical analyses were performed in STATA version 16·0, with weighting and clustering acknowledged using the complex survey functions.

## Results

9488 samples were included of which 8636 (91%) could be linked to a pseudo-identifier (Figure S1). These 8636 samples were derived from 1434 residents (2833 samples) and 3288 staff (5803 samples) (Table 1). A mean of 1.98 (SD 0·88) samples were collected from residents in 176 LTCFs and 1·76 (SD 0.79) samples from staff in 201 LTCFs. The median age was 48 years (IQR 35-56) in staff and 87 years (IQR 81-92) in residents. Across all LTCFs the mean proportion of residents sampled in at least one round of testing was 32% and 42% of staff, further details in Table S1.

**Table 1:** Demographics of study cohort.

|  | Proportion of samples that were antibody-positive* (%) | Proportion of individuals that were antibody-positive at any time* (%) |
| --- | --- | --- |
| Total | 2516 / 9488 (26.5) | 1361 / 4722 (28.5) |
| **Sex** |  |  |
| Female | 1895 / 7089 (26.7) | 1124 / 3892 (28.9) |
| Male | 424 / 1545 (27.4) | 237 / 828 (28.6) |
| Unknown | 143 / 854 (16.7) | 0 / 2 (0) |
| **LTCF role** |  |  |
| Resident | 965 / 3016 (32.0) | 484 / 1434 (33.8) |
| Staff member | 1551 / 6472 (16.7) | 877 / 3288 (26.7) |
| **Region** |  |  |
| London | 291 / 809 (36.0) | 155 / 350 (44.3) |
| South East | 473 / 1683 (28.1) | 259 / 861 (30.1) |
| East of England | 95 / 574 (16.6) | 47 / 261 (18.0) |
| East Midlands | 164 / 911 (18.0) | 96 / 521 (18.4) |
| West Midlands | 124 / 607 (20.4) | 73 / 318 (23.0) |
| South West | 328 / 1616 (20.3) | 184 / 873 (21.1) |
| North West | 297 / 1042 (28.5) | 168 / 538 (31.2) |
| North East | 590 / 1571 (37.6) | 290 / 654 (44.3) |
| Yorkshire & Humber | 154 / 675 (22.8) | 89 / 346 (25.7) |
| **LTCF Type** |  |  |
| For Profit chain | 1860 / 6503 (28.6) | 966 / 2989 (32.3) |
| Not for Profit chain | 563 / 2429 (23.2) | 320 / 1326 (24.1) |
| Independent | 93 / 506 (18.4) | 75 / 407 (18.4) |
| **LTCF size** |  |  |
| Small (<50 beds) | 916 / 3843 (23.8) | 492 / 1939 (25.4) |
| Medium (50-100 beds) | 1549 / 5491 (28.2) | 839 / 2688 (31.2) |
| Large (≥100 beds) | 51 / 154 (33.1) | 30 / 95 (31.6) |
| **Interval** |  |  |
| 1: June-July 2020 | 694 / 2225 (31.2) | NA |
| 2: August-September 2020 | 495 / 1794 (27.6) | NA |
| 3: October-November 2020 | 360 / 1349 (26.7) | NA |
| 4: December 2020-January 2021 | 200 / 1136 (17.6) | NA |
| 5: February 2021 | 221 / 920 (24.0) | NA |
| 6: March-April 2021 | 546 / 2064 (26.5) | NA |

\*Based on nucleocapsid antibody detected using Abbott assay
NA = Not Applicable

Antibodies to nucleocapsid were detected in 2516 (26·5%) samples, Table 1. Antibody-positivity varied by region and was greatest in London and the North East and lowest in East of England and the East Midlands. The proportion who were antibody-positive was highest in For-Profit chain and lowest in Independent LTCFs and varied over time with the lowest seroprevalence seen in December 2020 – January 2021. The proportion of antibody-positive individuals was lowest in LTCFs with less than 50 beds and highest in larger LTCFs, although only 154 samples were included from LTCFs with at least 100 beds.

The LTCF characteristics and the changes in LTCF seroprevalence are outlined in Table S1 and Figure S2.

### Weighted cumulative incidence and seroprevalence

Cumulative incidence in the whole population was 28·2% (95% CI 25·0-31·7%) and was significantly higher in residents 34·6% (95% 29·6-40·0) than in staff 26·1% (95% 23·0-29·5, p<0·0001), Figure 1, Table 2. Weighted seroprevalence was highest in both staff and residents in the first two months of testing (June-July 2020), just after the first wave of the pandemic compared with subsequent time intervals. Seroprevalence fell in staff over the following six months, reaching a minimum of 21.8% and 16.5% in residents and staff respectively in December 2020-January 2021 (interval 4), before rising again in March-April 2020 (interval 6) to 37·4% in residents and 23·0% in staff.

**Table 2:**
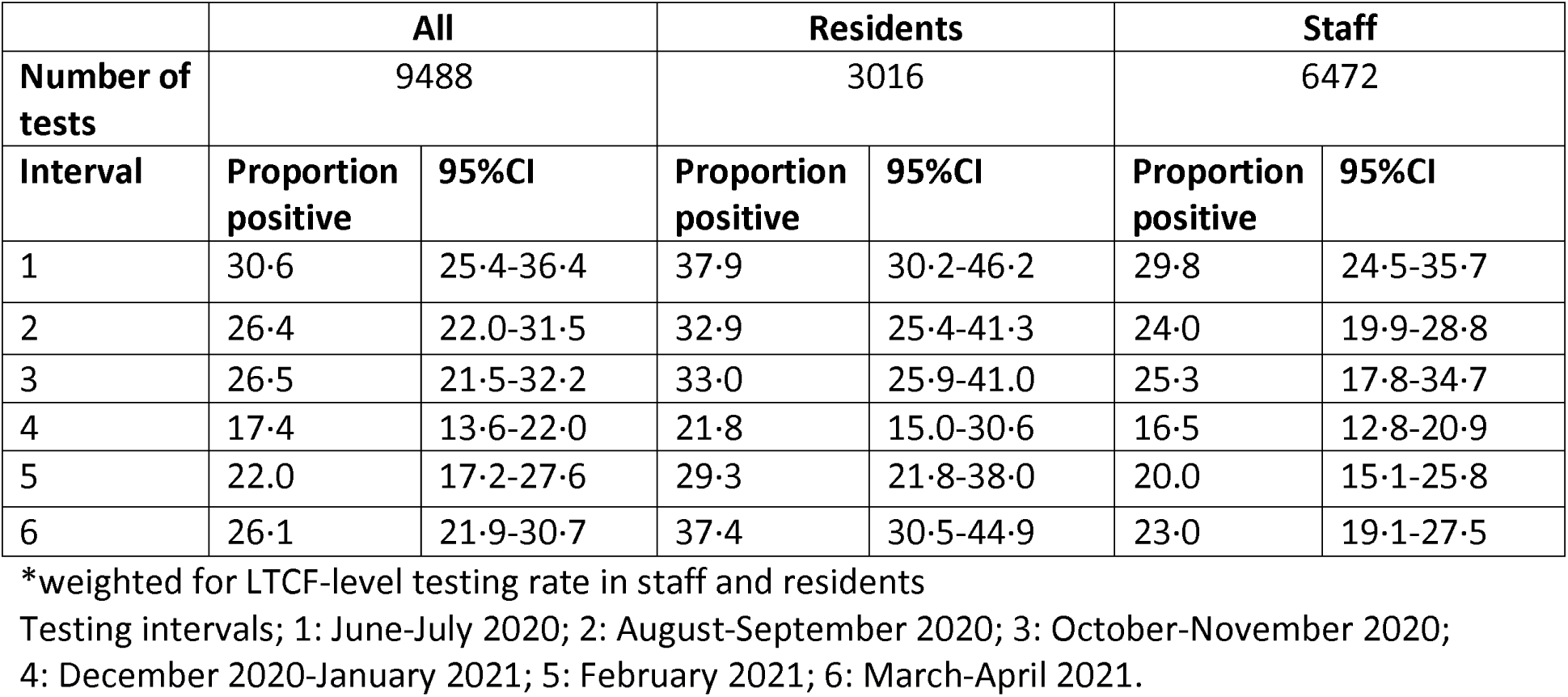
Weighted seroprevalence stratified by time interval and resident / staff role.

|  | **All** |  | **Residents** |  | **Staff** |  |
| --- | --- | --- | --- | --- | --- | --- |
| **Number of tests** | 9488 |  | 3016 |  | 6472 |  |
| **Interval** | **Proportion positive** | **95%CI** | **Proportion positive** | **95%CI** | **Proportion positive** | **95%CI** |
| 1 | 30.6 | 25.4-36.4 | 37.9 | 30.2-46.2 | 29.8 | 24.5-35.7 |
| 2 | 26.4 | 22.0-31.5 | 32.9 | 25.4-41.3 | 24.0 | 19.9-28.8 |
| 3 | 26.5 | 21.5-32.2 | 33.0 | 25.9-41.0 | 25.3 | 17.8-34.7 |
| 4 | 17.4 | 13.6-22.0 | 21.8 | 15.0-30.6 | 16.5 | 12.8-20.9 |
| 5 | 22.0 | 17.2-27.6 | 29.3 | 21.8-38.0 | 20.0 | 15.1-25.8 |
| 6 | 26.1 | 21.9-30.7 | 37.4 | 30.5-44.9 | 23.0 | 19.1-27.5 |

\*weighted for LTCF-level testing rate in staff and residents
Testing intervals; 1: June-July 2020; 2: August-September 2020; 3: October-November 2020; 4: December 2020-January 2021; 5: February 2021; 6: March-April 2021.

**Figure 1:**
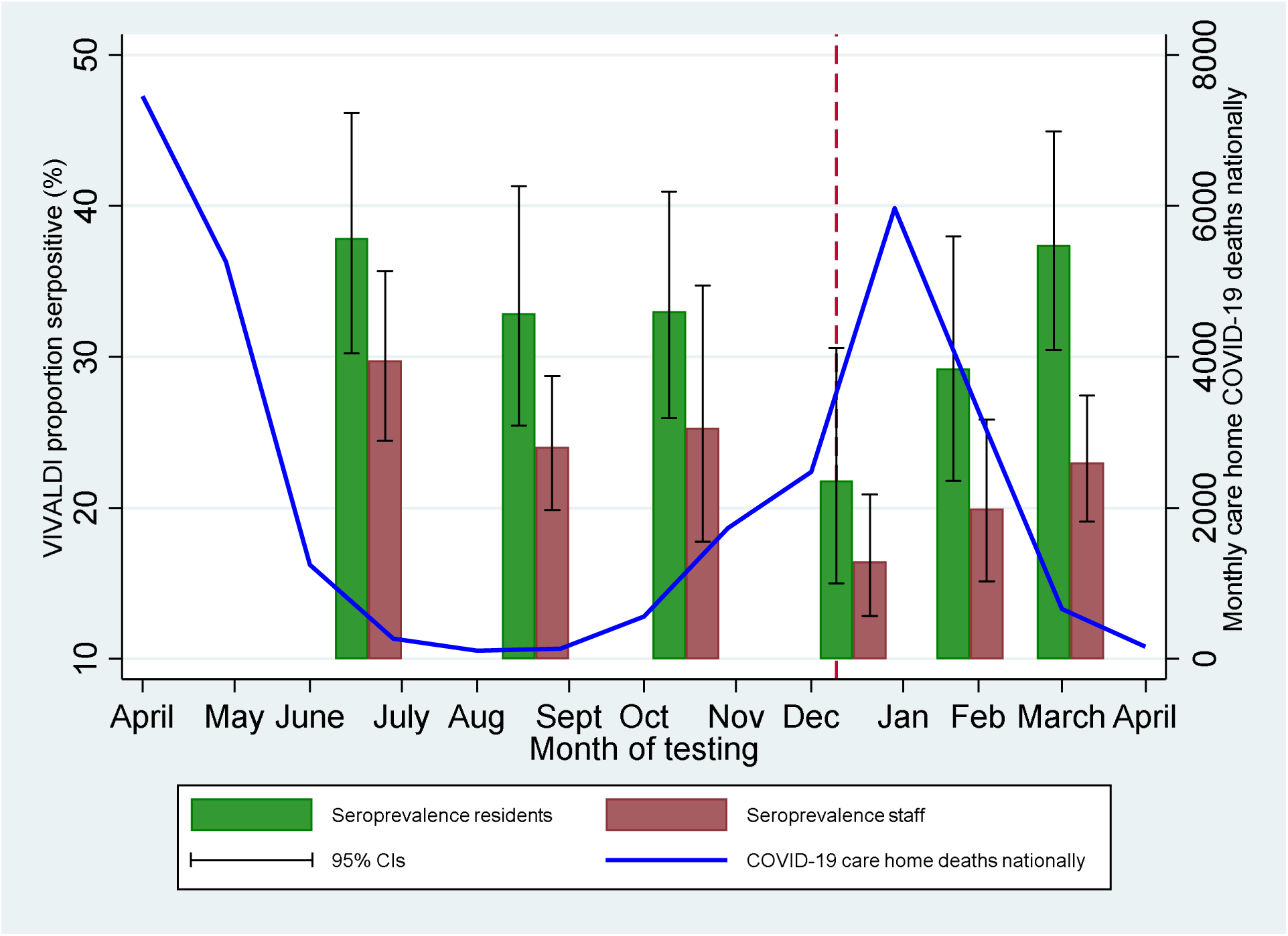
Weighted seroprevalence with 95% confidence intervals stratified by staff / resident and interval of testing compared against monthly COVID-19 associated deaths. COVID-19 associated deaths defined as deaths occurring within 28 days of COVID-19 diagnosis in LTCFs in England according to CQC reporting. ^30^ In view of limited PCR testing coverage in the first wave of the pandemic, data on COVID-19 deaths were considered a more accurate measure of the disease burden in LTCFs over the pandemic. *Red interrupted line represents start date of UK vaccination programme (8 December 2020)

### Time to antibody-loss

A total of 619 individuals were eligible for inclusion in the antibody-waning analysis (Table S2). This included 239 (38·6%) residents and 503 females (81·3%) with a median age of 59 years (IQR: 45-82 years). Further details on the included samples and antibody testing results by round are available in Tables S2-S4.

377 out of 380 staff had a positive baseline antibody sample and at least one subsequent sample, Tables S3a and S3b. Of these 380 staff, 218 were sampled in at least 2 subsequent testing rounds and 137 remained antibody-positive. 239 residents had a positive baseline antibody sample, of whom 154 had a third blood sample and 119 remained sero-positive. There were 3 cases of sero-conversion during follow-up and all occurred in staff.

Sero-reversion, defined as loss of a detectable nucleocapsid-specific antibody response,, occurred in 55 of 239 residents (23%) and 133 of 380 staff (35%). In the majority of cases (129/188) sero-reversion occurred 90-180 days following the estimated date of sero-conversion (Table S4).

Kaplan-Meier plots were used to estimate the median time to sero-reversion, Figure 2. The overall incidence rate of sero-reversion was 2·1 per 1000 person-days at risk and the estimated median time to sero-reversion was 242·5 days. The incidence was greater in staff compared to residents (2·4 vs 1·5 per 1000 person-days at risk, p=0·0003) (Figure 2, Table S5).

**Figure 2:**
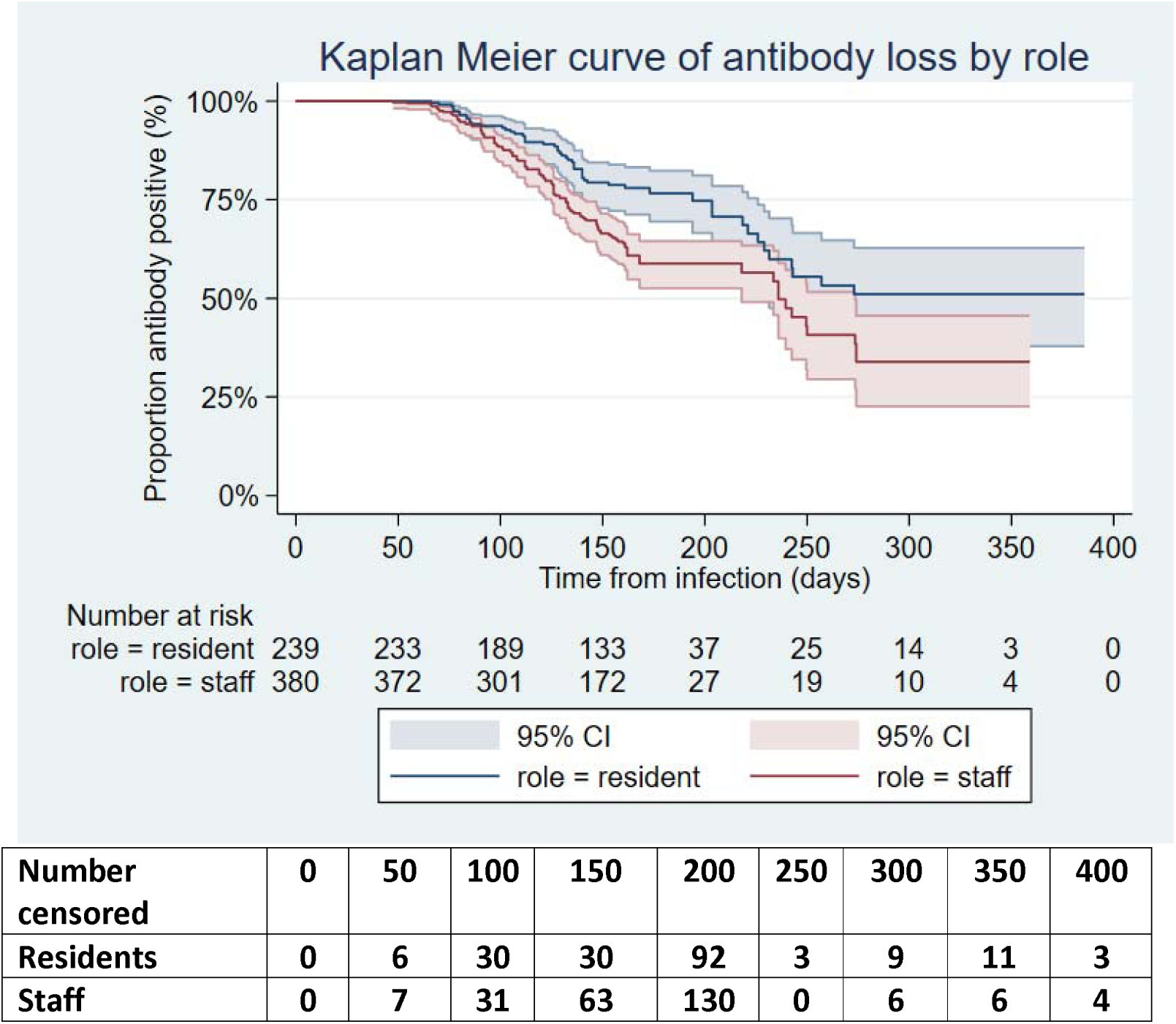
Kaplan Meier plot of time to antibody loss from estimated date of seroconversion in staff and residents with risk table and number censored (n=619)

Twenty individuals were admitted to hospital with confirmed or suspected COVID-19 prior to their first positive antibody test. Sub-group analysis estimated no significant difference in incidence rate of antibody-loss between hospitalised and non-hospitalised individuals (1·1 vs 2·1 per 1000 person-days at risk, p=0·1480) (figure S3 and table S6).

Quantitative antibody titres to spike and RBD proteins were plotted over time in a subset of 41 individuals (18 residents, 23 staff) who had sero-reverted. Samples were available at 60-90 days following sero-reversion for 16 individuals (7 staff, 9 residents). For all non-nucleocapsid antibodies tested, the median antibody titre did not recede below the threshold for positivity (figure 3a and b).

**Figure 3:**
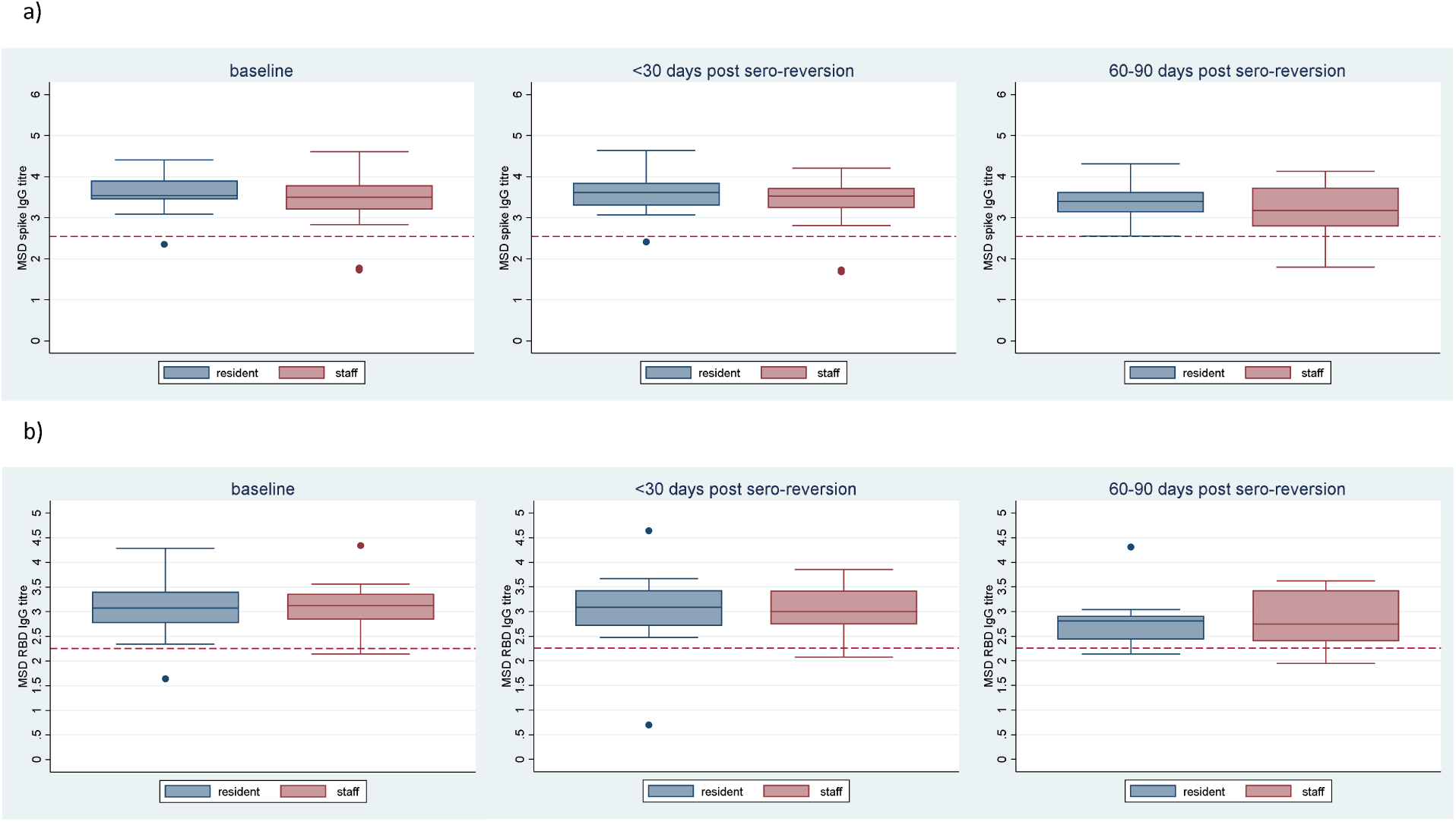
Quantitative antibody titres over 90 days following nucleocapsid antibody sero-reversion for a) spike antibody b) RBD antibody. Titres are presented at date of first positive antibody (baseline), T1: 0-30 days and T2: 60-90 days after estimated date of sero-reversion, (n=41 at baseline, and T1 and 16 at T2). Titres are reported from MSD assay according to a logarithmic scale. Red interrupted line denotes cut-off for test positivity (spike = 350 AU/ml, RBD = 180 AU/ml).

## Discussion

In the first two waves of the pandemic at least one-third of residents and one-quarter of staff in this study were infected with SARS-CoV-2. Seroprevalence was significantly greater in residents compared with staff and declined in the winter months between pandemic waves, which suggests significant waning of antibody levels. Indeed, around one half of the antibody-positive population sero-reverted within eight months based on antibodies to nucleocapsid although spike antibody titres remained elevated for at least an additional 90 days. A potential implication of these findings is that vaccine effectiveness in LTCF staff and residents in the context of high background prevalence of natural infection may differ from estimates of vaccine effectiveness in infection-naïve populations. This is because immunological studies suggest that natural infection augments vaccine-induced cellular and humoral immune responses in care home residents and staff.

In common with other serological studies, our study highlights the limitations of estimating the prevalence of natural infection based on antibody titres against nucleocapsid protein. ^25^ Direct comparison of seroprevalence estimates between studies and countries is challenging due to variation in the timing of pandemic waves and waning immunity. The largest published prevalence study to date included over 9,000 LTCF residents from 362 LTCFs in Madrid and reported that 55·4% of residents had detectable anti-nucleocapsid antibodies following the first wave of the pandemic (June to Nov 2020), however as these estimates were unweighted and the study was limited to one region, this may not be generalisable to the national prevalence of infection.^26^ In England, national surveillance data from 14 June-11 July 2021 report 16·6 % weighted nucleocapsid antibody prevalence in the general population, lowest in the 70-84 age group.^12^ These estimates are based on samples from blood donors in the community where infection rates were likely to be lower than in LTCFs, particularly over the first pandemic wave.^2^ Our study also found lower seroprevalence in Independent LTCFs when compared with For-Profit chains and in larger vs smaller LTCFs, which have been widely described as risk factors for SARS-CoV-2 transmission. ^1,27^

Our study makes a valuable contribution to a growing body of evidence of rapid waning of nucleocapsid antibody titres following natural infection across age groups, which presents challenges for measuring natural infection. ^19,25^ Although serological follow-up has been reported from LTCFs for up to four months, ^9^ we present antibody titres over eleven months and report that around one half of individuals will have undetectable nucleocapsid antibodies eight months from initial sero-conversion. One key factor relates to the sensitivity of the nucleocapsid-specific antibody assay that is used by each centre. The Abbott assay is used widely and has consistently been found to have sufficient diagnostic accuracy in head-to-head comparisons with other available assays. ^22,28^ Furthermore, to improve sensitivity whilst maintaining specificity we implemented a lower threshold for positivity (index value 0.8). ^22,23^

This study is one of the largest longitudinal sero-surveys that has been undertaken in frail older adults in LTCFs, who are often excluded from research studies. Participants were recruited from a range of small and large LTCFs including For-Profit and Not-for-Profit Providers in all regions in England, and longitudinal samples were obtained from >1500 frail residents. As we sampled a range of care providers and geographic regions in England, our results are likely to be generalisable to the wider LTCF population in this country. However, despite our efforts to use consultees for residents lacking capacity to consent, it is possible that this population are under-represented in our cohort due to the additional complexities that this posed for the LTCFs.

Despite a large sample, 13% of LTCFs did not sample any residents, highlighting the challenges of recruiting residents into research studies. We attempted to account for this, by stratifying our estimates by time interval and subject type and weighting for the LTCF size. Due to limited PCR and antibody testing nationally over the first wave of the pandemic, in some cases we estimated the date of primary infection and subsequent seroconversion using available data, which may have affected precision. Finally, we only sampled individuals who survived SARS-CoV-2 infection, which means we cannot infer the proportion of all care home residents (survivors and those who died) who were infected over successive waves of pandemic.

Although LTCFs were severely affected in the first wave of the pandemic, national infection rates have dropped significantly since the roll-out of vaccination. ^12^ Immunological studies have shown that prior natural infection with SARS-CoV-2 elicits a humoral immune response to COVID-19 vaccination that has a greater magnitude than that seen in infection-naïve people. ^5,6^ As a large proportion of people in LTCFs have had past infection, this may have overestimated the protective effect of vaccination in care home staff and residents. The median duration of residency in a LTCF is 462 days ^29^ and there is considerable staff turnover. As such it may be possible that lower vaccine effectiveness will be observed in LTCFs over time as the proportion of naturally infected individuals declines.

## Conclusion

Over one third of LTCF residents and one quarter of staff in England have evidence of SARS-CoV-2 infection over the first two waves of the pandemic. However, antibody titres against nucleocapsid protein wane significantly over the first eight months following infection. As this assay is the current gold-standard for assessment of prior natural infection it is likely that many people with previous natural infection will be falsely labelled as ‘infection-naïve’. Prior natural infection status is an important determinant of both the vaccine response and risk of reinfection and as such novel serological assays to determine status of prior infection should be developed.

## Data Availability

De-identified test results and limited meta-data will be made available for use by researchers in future studies, subject to appropriate research ethical approvals once the VIVALDI study cohort has been finalised. These datasets will be accessible via the Health Data Research UK Gateway.

## Contributors

LS, AH, AC, TP and MK conceptualised the study. MK, TP, LS and AC developed the statistical analysis plan. MK did the formal statistical analysis. MK, CF, BA, RG, MS and AI-S were involved with project administration. MK did the literature review. LS and AH obtained research funding. GT, NK, PS, TL and PM did laboratory investigations. MK wrote the first draft of the manuscript. All authors revised and edited the manuscript. MK and TP accessed and verified the data. All authors had full access to the all the data reported in the study. LS, AC and MK shared the final responsibility for the decision to submit for publication.

## Acknowledgments

We thank the staff and residents in the LTCFs that participated in this study, Rachel Bruton who project managed the study at the University of Birmingham, and Mark Marshall at NHS England who pseudonymised the electronic health records. This report is independent research funded by the Department of Health and Social Care (COVID-19 surveillance studies). MK is funded by a Wellcome Trust Clinical PhD Fellowship (222907/Z/21/Z). LS is funded by a National Institute for Health Research Clinician Scientist Award (CS-2016-007). AH is supported by Health Data Research UK (LOND1), which is funded by the UK Medical Research Council, Engineering and Physical Sciences Research Council, Economic and Social Research Council, Department of Health and Social Care (England), Chief Scientist Office of the Scottish Government Health and Social Care Directorates, Health and Social Care Research and Development Division (Welsh Government), Public Health Agency (Northern Ireland), British Heart Foundation, and Wellcome Trust. The views expressed in this publication are those of the authors and not necessarily those of the NHS, Public Health England, or the Department of Health and Social Care.

## References

1 Burton JK, Bayne G, Evans C, et al. Evolution and effects of COVID-19 outbreaks in care homes: a population analysis in 189 care homes in one geographical region of the UK. The Lancet Healthy Longevity 2020; 1: e21–31.

2 Lemmon E. http://ltccovid.org | COVID-19 mortality and long-term care: a UK comparison COVID-19 mortality and long-term care: a UK comparison. 2020. https://ltccovid.org/wp-content/uploads/2020/08/COVID-19-mortality-in-long-term-care-final-Sat-29-v1.pdf (accessed July 12, 2021).

3 Long Q-X, Liu B-Z, Deng H-J, et al. Antibody responses to SARS-CoV-2 in patients with COVID-19. Nature Medicine 2020; : 1–4.

4 Huang AT, Garcia-Carreras B, Hitchings MDT, et al. A systematic review of antibody mediated immunity to coronaviruses: kinetics, correlates of protection, and association with severity. Nature Communications 2020; 11. DOI:10.1038/S41467-020-18450-4.

5 Krammer F, Srivastava K, Alshammary H, et al. Antibody Responses in Seropositive Persons after a Single Dose of SARS-CoV-2 mRNA Vaccine. New England Journal of Medicine 2021; 384: 1372–4.

6 Tut G, Lancaster T, Krutikov M, et al. Profile of humoral and cellular immune responses to single doses of BNT162b2 or ChAdOx1 nCoV-19 vaccines in residents and staff within residential care homes (VIVALDI): an observational study. The Lancet Healthy Longevity 2021; 2: e544–53.

7 Ebinger JE, Fert-Bober J, Printsev I, et al. Antibody responses to the BNT162b2 mRNA vaccine in individuals previously infected with SARS-CoV-2. Nature Medicine 2021; 27: 981–4.

8 Jeffery-Smith A, Dun-Campbell K, Janarthanan R, et al. Infection and transmission of SARS-CoV-2 in London care homes reporting no cases or outbreaks of COVID-19: Prospective observational cohort study, England 2020. The Lancet Regional Health - Europe 2021; 3: 100038.

9 Jeffery-Smith A, Iyanger N, Williams S v., et al. Antibodies to SARS-CoV-2 protect against re-infection during outbreaks in care homes, September and October 2020. Eurosurveillance 2021; 26: 2100092.

10 Krutikov M, Palmer T, Tut G, et al. Incidence of SARS-CoV-2 infection according to baseline antibody status in staff and residents of 100 long-term care facilities (VIVALDI): a prospective cohort study. The Lancet Healthy Longevity 2021; 2: e362–70.

11 Office for National Statistics. Coronavirus (COVID-19) Infection Survey technical article. 2021 https://www.ons.gov.uk/peoplepopulationandcommunity/healthandsocialcare/conditionsanddiseases/articles/coronaviruscovid19infectionsurveytechnicalarticle/wavesandlagsofcovid19inenglandjune2021 (accessed July 27, 2021).

12 Public Health England. Weekly national Influenza and COVID-19 surveillance report Executive summary - Week 29 2021. 2021 https://assets.publishing.service.gov.uk/government/uploads/system/uploads/attachment_data/file/1005056/Weekly_Flu_and_COVID-19_report_w29.pdf (accessed Aug 17, 2021).

13 GOV.UK. Vaccine update: issue 315, December 2020, COVID-19 special edition - GOV.UK. https://www.gov.uk/government/publications/vaccine-update-issue-315-december-2020-covid-19-special-edition/vaccine-update-issue-315-december-2020-covid-19-special-edition#covid-19-vaccination-programme (accessed July 12, 2021).

14 Blain H, Tuaillon E, Gamon L, et al. Spike Antibody Levels of Nursing Home Residents with or without Prior COVID-19 3 Weeks after a Single BNT162b2 Vaccine Dose. JAMA - Journal of the American Medical Association. 2021; 325: 1898–9.

15 Houlihan CF, Beale R. The complexities of SARS-CoV-2 serology. The Lancet Infectious Diseases. 2020; 20: 1350–1.

16 Seow J, Graham C, Merrick B, et al. Longitudinal observation and decline of neutralizing antibody responses in the three months following SARS-CoV-2 infection in humans. Nature Microbiology 2020; 5: 1598–607.

17 Ibarrondo FJ, Fulcher JA, Goodman-Meza D, et al. Rapid Decay of Anti–SARS-CoV-2 Antibodies in Persons with Mild Covid-19. New England Journal of Medicine 2020; 383: 1085–7.

18 van Elslande J, Oyaert M, Ailliet S, et al. Longitudinal follow-up of IgG anti-nucleocapsid antibodies in SARS-CoV-2 infected patients up to eight months after infection. Journal of Clinical Virology 2021; 136: 104765.

19 Alfego D, Sullivan A, Poirier B, Williams J, Adcock D, Letovsky S. A population-based analysis of the longevity of SARS-CoV-2 antibody seropositivity in the United States. EClinicalMedicine 2021; 36: 100902.

20 Krutikov M, Palmer T, Donaldson A, et al. Study Protocol: Understanding SARS-Cov-2 infection, immunity and its duration in care home residents and staff in England (VIVALDI). Wellcome Open Research 2020; 5: 232.

21 Department of Health & Social Care. Coronavirus (COVID-19) Scaling up our testing programmes Department of Health and Social Care. 2020 https://assets.publishing.service.gov.uk/government/uploads/system/uploads/attachment_data /file/878121/coronavirus-covid-19-testing-strategy.pdf (accessed Dec 22, 2020).

22 Ainsworth M, Andersson M, Auckland K, et al. Performance characteristics of five immunoassays for SARS-CoV-2: a head-to-head benchmark comparison. The Lancet Infectious Diseases 2020; 20: 1390–400.

23 Bryan A, Pepper G, Wener M, et al. Performance Characteristics of the Abbott Architect SARS-CoV-2 IgG Assay and Seroprevalence in Boise, Idaho. Journal of clinical microbiology 2020; 58. DOI:10.1128/JCM.00941-20.

24 Dowell AC, Butler MS, Jinks E, et al. Children develop strong and sustained cross-reactive immune responses against Spike protein following SARS-CoV-2 infection, with enhanced recognition of variants of concern. medRxiv 2021; : 2021.04.12.21255275.

25 Ward H, Cooke GS, Atchison C, et al. Prevalence of antibody positivity to SARS-CoV-2 following the first peak of infection in England: Serial cross-sectional studies of 365,000 adults. The Lancet Regional Health - Europe 2021; 4: 100098.

26 Candel FJ, Barreiro P, San Román J, et al. The demography and characteristics of SARS-CoV-2 seropositive residents and staff of nursing homes for older adults in the Community of Madrid: the SeroSOS study. Age and Ageing 2021; 50: 1038–47.

27 Stall NM, Jones A, Brown KA, Rochon PA, Costa AP. For-profit long-term care homes and the risk of COVID-19 outbreaks and resident deaths. CMAJL: Canadian Medical Association Journal 2020; 192: E946.

28 Patel EU, Bloch EM, Clarke W, et al. Comparative performance of five commercially available serologic assays to detect antibodies to SARS-CoV-2 and identify individuals with high neutralizing titers. Journal of Clinical Microbiology 2021; 59. DOI:10.1128/JCM.02257-20.

29 Forder J, Fernandez J-L. Length of stay in care homes. 2011 www.pssru.ac.uk (accessed July 12, 2021).

30 Office for National Statistics. Number of deaths in care homes notified to the Care Quality Commission, England. 2021. https://www.ons.gov.uk/peoplepopulationandcommunity/birthsdeathsandmarriages/deaths/datasets/numberofdeathsincarehomesnotifiedtothecarequalitycommissionengland (accessed July 12, 2021).

