## Supplementary appendix for "Prevalence and duration of detectable SARS-CoV-2 nucleocapsid antibody in staff and residents of long-term care facilities over the first year of the pandemic (VIVALDI study): prospective cohort study"

**Figure S1: study inclusion flow diagram**

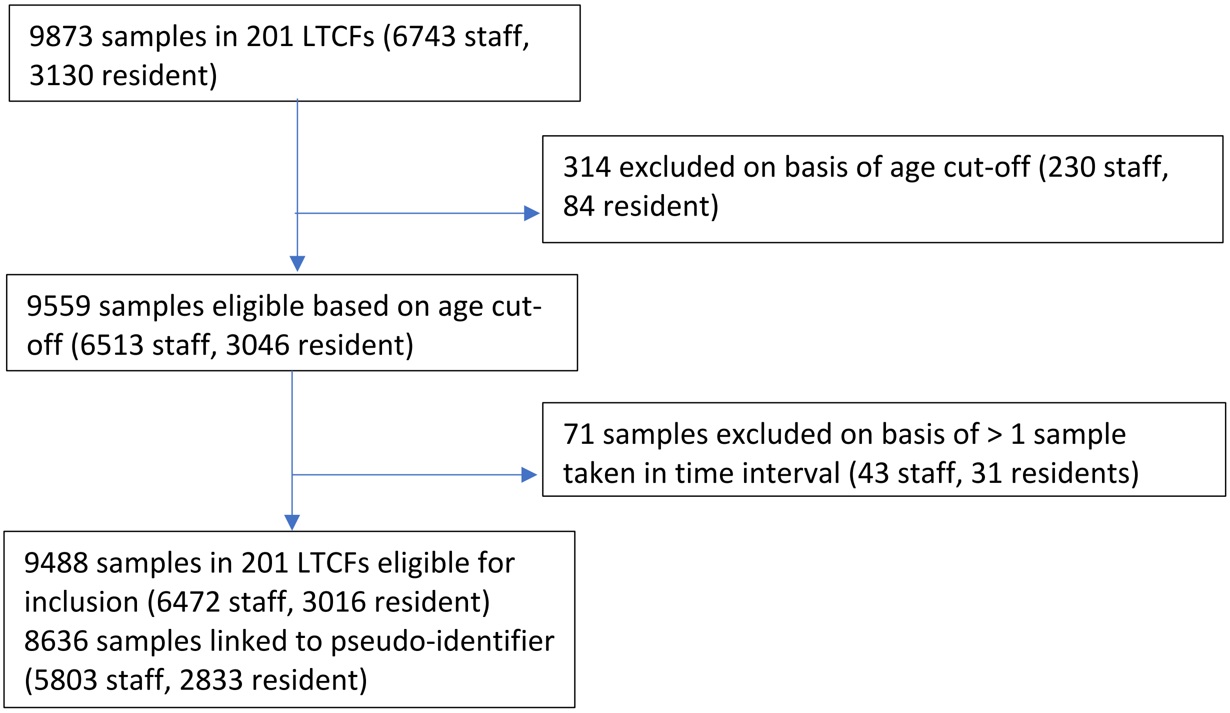

**Figure S2: Changes in LTCF weighted seroprevalence over successive sampling rounds according to 'baseline' LTCF seroprevalence in round 1 (n=201)**

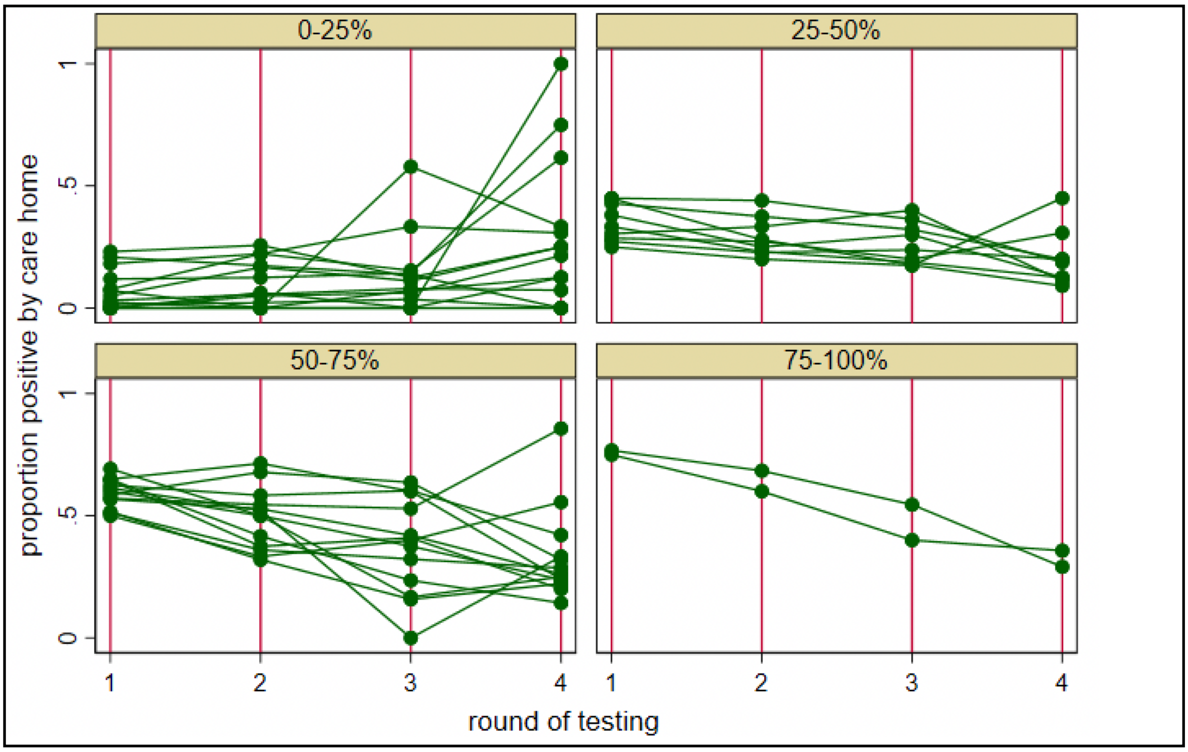

**Figure S3: Kaplan Meier plot of time to antibody loss from estimated date of seroconversion according to severity of primary infection (hospital admission vs no hospital admission) in person-days**

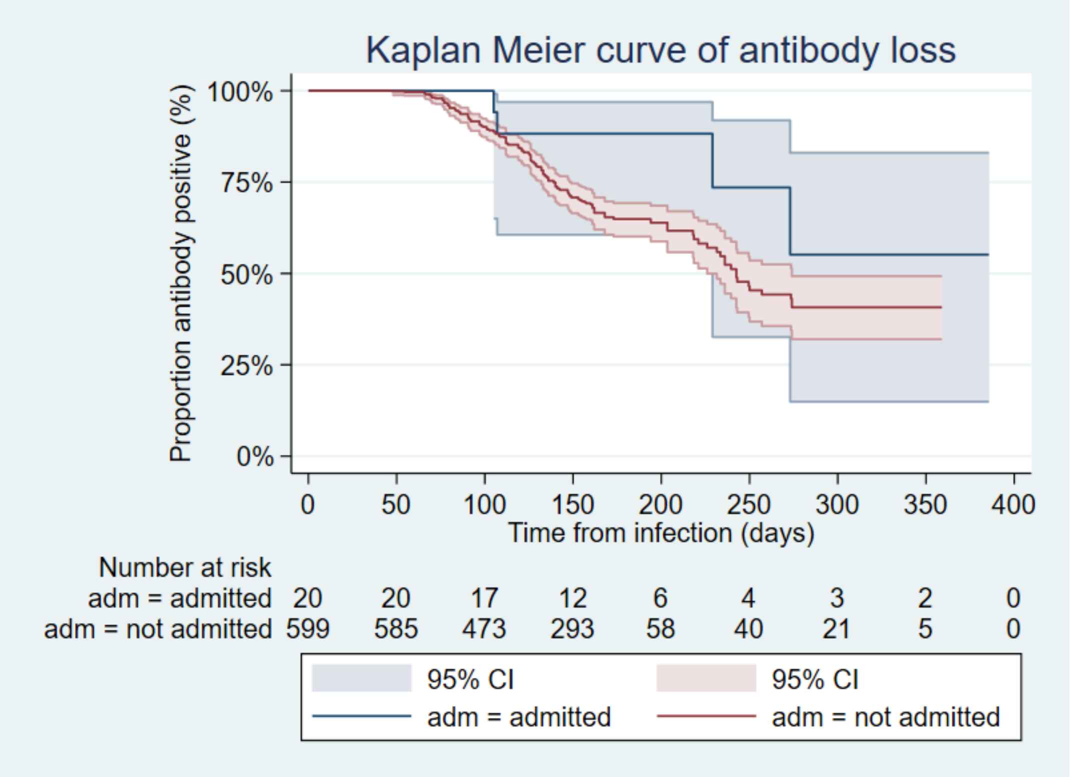

| Number censored | 0 | 50 | 100 | 150 | 200 | 250 | 300 | 350 | 400 |
|---|---|---|---|---|---|---|---|---|---|
| Admitted | 0 | 0 | 3 | 3 | 6 | 1 | 0 | 1 | 2 |
| Not admitted | 0 | 13 | 58 | 90 | 216 | 2 | 15 | 16 | 5 |

### Table S1: Characteristics of included LTCFs

| | **Number of LTCFs (%)** |
|---|---|
| Total | 201 |
| Proportion where residents sampled | 176 (87·6) |
| Proportion where staff sampled | 201 (100) |
| Region | |
| London | 10 (5·0) |
| South East | 40 (19·9) |
| East of England | 13 (6·5) |
| East Midlands | 26 (12·9) |
| West Midlands | 10 (5·0) |
| South West | 39 (19·4) |
| North West | 29 (14·4) |
| North East | 20 (10·0) |
| Yorkshire & Humber | 14 (7·0) |
| LTCF type | |
| For-profit chain | 118 (58·7) |
| Not for Profit chain | 64 (31·8) |
| Independent | 19 (9·5) |
| Round of testing | |
| 1 | 201 (100) |
| 2 | 175 (87·1) |
| 3 | 84 (41·8) |
| 4 | 39 (19·4) |
| Interval | |
| 1: June-July 2020 | 96 (19·6) |
| 2: August-September 2020 | 94 (19·2) |
| 3. October-November 2020 | 87 (17·8) |
| 4. December 2020-January 2021 | 53 (10·8) |
| 5. February 2021 | 59 (12·0) |
| 6. March-April 2021 | 101 (20·6) |
| Occupied beds per LTCF, mean (SD) | 44·36 (16·5) |
| Number of staff per LTCF, mean (SD) | 56·86 (21·9) |
| Number of samples per LTCF per round, mean (SD): | |
| Staff | 13·18 (8·61) |
| Residents | 7·60 (6·05) |

### Table S2: Number of samples included in the analysis by care home role and mean time between samples in days

| **Testing round** | **Number of Residents with antibodies to N (%)** | **Number of Staff with antibodies to N (%)** | **Mean time to next sample in days (SD)** |
|---|---|---|---|
| **1** | 239/239 (100) | 377/380 (99·2) | 62·5 (26·9) |
| **2** | 211/239 (88·3) | 303/380 (79·7) | 61·8 (24·0) |
| **3** | 119/154 (77·3) | 140/218 (64·2) | 157·2 (24·9) |
| **4** | 23/37 (62·2) | 8/18 (44·4) | NA |

NA = Not Applicable

**Table S3: Antibody results by round of testing and round 1 antibody result in a) staff b) residents**

**a)**

| | | Round 2 | | | Round 3 | | | Round 4 | | |
|---|---|---|---|---|---|---|---|---|---|---|
| | | Positive (%) | Negative (%) | Total | Positive (%) | Negative (%) | Total | Positive (%) | Negative (%) | Total |
| Round 1 | Positive (%) | 300 (79·6) | 77 (20·4) | 377 | 137 (63·7) | 78 (36·3) | 215 | 7 (41·2) | 10 (58·8) | 17 |
| | Negative (%) | 3 (100) | 0 (0) | 3 | 3 (100) | 0 (0) | 3 | 1 (100) | 0 (0) | 1 |
| Total | | 303 | 77 | 380 | 140 | 78 | 218 | 8 | 10 | 18 |

**b)**

| | | Round 2 | | | Round 3 | | | Round 4 | | |
|---|---|---|---|---|---|---|---|---|---|---|
| | | Positive (%) | Negative (%) | Total | Positive (%) | Negative (%) | Total | Positive (%) | Negative (%) | Total |
| Round 1 | Positive (%) | 211 (88·3) | 28 (11·7) | 239 | 119 (77·3) | 35 (22·7) | 154 | 23 (62·2) | 14 (37·8) | 37 |
| | Negative (%) | 0 | 0 | 0 | 0 | 0 | 0 | 0 | 0 | 0 |
| Total | | 211 | 28 | 239 | 119 | 35 | 154 | 23 | 14 | 37 |

**Table S4: Distribution of time to sero-reversion for residents and staff (n=188)***

| Time to sero-reversion | Residents (%) | Staff (%) | Total (%) |
|---|---|---|---|
| < 90 days | 13 (23·6) | 23 (17·3) | 36 (19·2) |
| 90-180 days | 30 (54·6) | 99 (74·4) | 129 (68·6) |
| 180-270 days | 11 (20·0) | 8 (6·0) | 19 (10·1) |
| ≥ 270 days | 1 (1·8) | 3 (2·3) | 4 (2·1) |
| Total | 55 | 133 | 188 |

* Among those that sero-reverted, those who did not (censored) are excluded

**Table S5: Time at risk and incidence rate of sero-reversion by care home role in person-days**

| Role | Time at risk (person-days) | Incidence rate (per 1000 person-days) |
|---|---|---|
| Staff | 54543 | 2·44 |
| Residents | 37141 | 1·48 |
| Overall | 91684 | 2·05 |

**Table S6: Time at risk and incidence rate according to severity of primary infection**

| Severity of infection | Time at risk (person-days) | Incidence rate (per 1000 person-days) |
|---|---|---|
| Admitted to hospital | 3639 | 1·10 |
| Not admitted to hospital | 88045 | 2·09 |

### STROBE checklist

| | Item No | Recommendation | Page No |
|---|---|---|---|
| Title and abstract | | | |
| | 1 | (*a*) Indicate the study's design with a commonly used term in the title or the abstract | 1 |
| | | (*b*) Provide in the abstract an informative and balanced summary of what was done and what was found | 2 |
| Introduction | | | |
| Background/rationale | 2 | Explain the scientific background and rationale for the investigation being reported | 4 |
| Objectives | 3 | State specific objectives, including any prespecified hypotheses | 4 |
| Methods | | | |
| Study design | 4 | Present key elements of study design early in the paper | 4-5 |
| Setting | 5 | Describe the setting, locations, and relevant dates, including periods of recruitment, exposure, follow-up, and data collection | 4-5 |
| Participants | 6 | *a) Cohort study*? Give the eligibility criteria, and the sources and methods of selection of participants. Describe methods of follow-up<br>*Case-control study*? Give the eligibility criteria, and the sources and methods of case ascertainment and control selection. Give the rationale for the choice of cases and controls<br>*Cross sectional study*? Give the eligibility criteria, and the sources and methods of selection of participants | 4-6 |
| | | (*b*) *Cohort study*? For matched studies, give matching criteria and number of exposed and unexposed<br>*Case-control study*? For matched studies, give matching criteria and the number of controls per case | n/a |
| Variables | 7 | Clearly define all outcomes, exposures, predictors, potential confounders, and effect modifiers. Give diagnostic criteria, if applicable | 5-7 |
| Data sources/ measurement | 8* | For each variable of interest, give sources of data and details of methods of assessment (measurement). Describe comparability of assessment methods if there is more than one group | 5-7 |
| Bias | 9 | Describe any efforts to address potential sources of bias | 4-7 |
| Study size | 10 | Explain how the study size was arrived at | 7 |
| Quantitative variables | 11 | Explain how quantitative variables were handled in the analyses. If applicable, describe which groupings were chosen and why | 6-7 |
| Statistical methods | 12 | (*a*) Describe all statistical methods, including those used to control for confounding | 6-7 |
| | | (*b*) Describe any methods used to examine subgroups and interactions | 6-7 |
| | | (*c*) Explain how missing data were addressed | 5-7 |
| | | (*d*) *Cohort study*? If applicable, explain how loss to follow-up was addressed<br>*Case-control study*? If applicable, explain how matching of cases and controls was addressed<br>*Cross sectional study*? If applicable, describe analytical methods taking account of sampling strategy | 5-7 |
| | | (*e*) Describe any sensitivity analyses | n/a |
| Results | | | |
| Participants | 13* | (*a*) Report numbers of individuals at each stage of study? eg numbers potentially eligible, examined for eligibility, confirmed eligible, included in the study, completing follow-up, and analysed | 7-9, figure S1 |
| | | (*b*) Give reasons for non-participation at each stage | 7-9 Figure S1 |

| | Item No | Recommendation | Page No |
|---|---|---|---|
| | | (*c*) Consider use of a flow diagram | Figure S1 (appendix) |
| Descriptive data | 14* | (*a*)Give characteristics of study participants (eg demographic, clinical, social) and information on exposures and potential confounders | 7-9, table 1, appendix |
| | | (*b*) Indicate number of participants with missing data for each variable of interest | 7-9 |
| | | (*c*) *Cohort study*? Summarise follow-up time (eg average and total amount) | 7-9 |
| Outcome data | 15* | *Cohort study*? Report numbers of outcome events or summary measures over time | 7-9, table 1-2, appendix |
| | | *Case-control study?* Report numbers in each exposure category, or summary measures of exposure | |
| | | *Cross sectional study?* Report numbers of outcome events or summary measures | |
| Main results | 16 | (*a*) Report the numbers of individuals at each stage of the study?eg numbers potentially eligible, examined for eligibility, confirmed eligible, included in the study, completing follow-up, and analysed | 7-9, appendix, table1-2 |
| | | (*b*) Give reasons for non-participation at each stage | 7-9, appendix |
| | | (*c*) Consider use of a flow diagram | |
| Other analyses | 17 | Report other analyses done?eg analyses of subgroups and interactions, and sensitivity analyses | 8-9, appendix |
| Discussion | | | |
| Key results | 18 | Summarise key results with reference to study objectives | 9-10 |
| Limitations | 19 | Discuss limitations of the study, taking into account sources of potential bias or imprecision. Discuss both direction and magnitude of any potential bias | 9-10 |
| Interpretation | 20 | Give a cautious overall interpretation of results considering objectives, limitations, multiplicity of analyses, results from similar studies, and other relevant evidence | 9-10 |
| Generalisability | 21 | Discuss the generalisability (external validity) of the study results | 9-10 |
| Other information | | | |
| Funding | 22 | Give the source of funding and the role of the funders for the present study and, if applicable, for the original study on which the present article is based | 7 |
